# Fluoroscopy time as a new predictor of short-term outcomes after transcatheter aortic valve replacement

**DOI:** 10.1101/2023.07.27.23293294

**Authors:** Alessandro Cafaro, Francesco Spione, Osvaldo Burattini, Daniele De Feo, Alessandro Xhelo, Chiara Palmitessa, Maurizio D’Alessandro, Vincenzo Pio Amendola, Flavio Rimmaudo, Andrea Igoren Guaricci, Alessandro Santo Bortone, Vincenzo Pestrichella, Gaetano Contegiacomo, Tullio Tesorio, Giuseppe Colonna, Fortunato Iacovelli

## Abstract

**Objective:** Transcatheter aortic valve replacement (TAVR) is a cine-fluoroscopic guided procedure. The amount of radiation used during the procedure is strictly related with fluoroscopy time (FT) that has been demonstrated to be associated with outcomes and complexity of procedure in percutaneous coronary interventions. The aim of our study is to demonstrate the relationship between FT and short-term outcomes after TAVR.

**Methods:** After splitting 1797 consecutive patients according to tertiles of FT, the composite endpoint early safety (ES) was adjudicated according to Valve Academic Research Consortium (VARC)-2 and -3 consensus documents and the composite endpoints of device success (DS) and technical success (TS) according to VARC-3 criteria.

**Results:** The absence of all outcomes (TS, DS, and ES according to VARC-3 and ES according to VARC-2) was significantly associated with higher FT and this association persisted after propensity score matching analysis. Notwithstanding, after receiver operating characteristic analysis, only the FT cut-offs identified for VARC-3 TS and VARC-2 ES had adequate diagnostic accuracy in identifying the absence of these endpoints.

**Conclusions:** Longer FT is related with peri-procedural and short-term outcomes after the procedure, especially in those that are more complex. A FT duration of more than 30 minutes has an adequate accuracy in identifying VARC-3 technical failure and absence of VARC-2 ES.

**What is already known on this topic:** FT is related with complexity and outcomes in PCI. No data is available about FT and TAVR.

**What this study adds:** FT >30 minutes has an adequate accuracy in identifying VARC-3 technical failure and absence of VARC-2 ES.

**How this study might affect research, practice or policy:** FT that lasts more than 30 minutes in TAVR is linked independently to short-term adverse outcomes after TAVR. A strict follow-up is needed in this procedural setting as FT is a new independent predictor of adverse outcome after TAVR.

## 1. Introduction

Transcatheter aortic valve replacement (TAVR) is an established treatment for pa-tients with aortic valve stenosis at high surgical risk or considered inadequate for con-ventional surgical aortic valve replacement (SAVR). Multiple observational and random-ized clinical trials have demonstrated the feasibility and efficacy of this treatment [1-10]. Notwithstanding, recent randomized trials have demonstrated that this percutaneous technique is non-inferior to SAVR also in intermediate and low surgical risk patients [11-15].

TAVR is a cine-fluoroscopic guided procedure, and the amount of radiation used is potentially dangerous for both operators and patients because of its stochastic and de-terministic adverse effects [16,17]. The radiation dose, which is strictly related to fluoros-copy time and procedure length, has been demonstrated to be similar to other percuta-neous coronary interventions of moderate complexity [18-20].

To date, no study has investigated the association between FT and short-term prog-nosis after TAVR. In particular, in the Valve Academic Research Consortium (VARC) documents [21,22], early safety (ES) and device success (DS) are short-term composite endpoints. ES combines all-cause mortality, all stroke, life-threatening bleeding, stage 2 or 3 acute kidney injury (AKI), coronary artery obstruction requiring intervention, and valve-related dysfunction requiring another aortic valvular procedure within 30 days after TAVR. DS combines the absence of procedural mortality, correct positioning of a single prosthetic heart valve into the proper anatomical location and intended performance of the prosthetic heart valve. The VARC-3 consensus document added the endpoint technical success (TS) which is a composite of freedom from mortality, successful access, delivery of the device, and retrieval of the delivery system, correct positioning of a single prosthetic heart valve into the proper anatomical location and freedom from surgery or intervention related to the devices or to a major vascular or access-related, or cardiac structural complications at the exit from procedure room [22]. With respect to ES definition, the VARC-3 document added other adverse events that significantly impact short-and long-term prognosis, such as cardiac structural complications, significant (moderate to severe) aortic regurgitation, and new permanent pacemaker (PM) implantation [22]. Finally, the definition of DS in the VARC-3 document added TS among the other endopoints included in the VARC-2 definition [21,22].

Our study aims to evaluate for the first time, in a large population, the relationship between FT and short-term outcomes after TAVR.

### 2. Materials and Methods

## 2.1. Study population

This multicenter observational study assessed all consecutive patients who under-went TAVR at five southern Italy heart centers (“Montevergine” Clinic of Mercogliano, Policlinico University Hospital, “Anthea” Clinic and “Mater Dei” Hospital of Bari and “V. Fazzi” Hospital of Lecce) involved in the “Magna Graecia” TAVR registry.

Between March 2011 and April 2023, 1797 consecutive patients (785 males, mean age 80.86±5.71, 1703 transfemoral access) suitable for TAVR were enrolled. All patients underwent preprocedural assessment with transthoracic echocardiography, coronary angiography, computed tomography scan of the heart, aorta and peripheral arteries, carotid artery ultrasonography and multidisciplinary evaluation by the Heart Team. The majority of procedures were performed in a standard cardiac catheterization laboratory with the support of anesthesiology and surgical backup by experienced operators. Device used were balloon-expandable valves (Edwards Sapien XT and Sapien 3; Meril Myval), self-expandable valves (Medtronic CoreValve, Engager, Evolut R and Evolut PRO; Abbott Portico) and others (Boston Lotus; Boston Acurate and Acurate neo; Direct Flow Medical; JenaValve).

Each participating site collected all baseline demographics, clinical, laboratory, echocardiographic, surgical risk score, intraprocedural and postprocedural data, in-hospital outcomes and 1-month follow-up outcomes, in the same dedicated archiving software. All the adverse events and the TS, DS and ES composite endpoints were also re-adjudicated retrospectively, by an external committee of interventional cardiologists, according to both VARC-2 and VARC-3 criteria [21,22]. All TAVR-related complications (according VARC-2 and VARC-3 definition both separately and then globally considered) were divided by intra-and post-procedural. Time delay between the end of TAVR procedure and the first post-procedural complication occurrence was also registered.

Patients’ population was retrospectively divided according to FT (minutes) tertiles and than by enrollment-time tertiles in order to study FT and radiation dose (RD) during TAVR learning curves.

## 2.2. Statistical analysis

Statistical analysis was performed using SigmaStat 3.5, SPSS 25.0 and STATA 13.0 softwares. Continuous variables were expressed as the mean ± standard deviation and median (interquartile ranges) of absolute numbers; categorical variables were expressed as frequencies and percentages. As appropriate, comparisons were made by t-test, Mann Whitney’s U-test, one way ANOVA, ANOVA on ranks, Fisher’s exact test or χ2 test. Pairwise multiple comparisons after ANOVAs were conducted through Holm-Sidak or Dunn’s test as properly indicated by definitions. The normal distribution was assessed with Kolmogo-rov-Smirnov tests. A receiver-operating characteristic (ROC) curve analysis was built in order to establish the threshold levels of FT that provided the best cut-off for the absence of ES according to VARC-2 and VARC-3 definitions. Area under the curve (AUC) values were calculated with confidence intervals (CIs) through concordance statistics to measure test accuracy. The DeLong test was used to identify AUC standard errors. The calibration of FT was evaluated by comparing the mean predicted probability and the mean observed frequency of absence of ES with goodness-of-fit R-squared and Cochran-Armitage tests, calibration plots and estimation of a calibration slope. After this, new optimal cut-off points for the absence of ES were selected using Youden’s tests, reporting Youden’s indexes: we evaluated sensitivity and specificity according to these new cut-off points. The relationship between FT and the absence of ES was also analysed after propensity score matching (PSM) including as covariates those factors that were considered to increase the time of the procedure: pre-dilatation, post-dilatation, intra-procedural complications, self expandable valve implantation, pre-TAVR ejection-fraction, pre-TAVR maximum transaortic gradient, trans-apical and direct aortic access. All statistical tests were two-sided. For all tests, a p-value <0.05 was considered statistically significant.

### 3. Results

## 3.1. Baseline Characteristics

Patients’ population was divided according to FT (minutes) tertiles: 1st group 13.94±2.93 min, 2nd group 21.31±1.99 min and 3rd group 38.31±18.83 min.

All clinical and preprocedural data of the study population are shown in Table 1.

**Table 1.** Baseline characteristics of the study population according to FT tertiles.

| Variable | All | Fluoroscopy Time |  |  |  |
| --- | --- | --- | --- | --- | --- |
|  |  | 1<sup>st</sup> | 2<sup>nd</sup> | 3<sup>rd</sup> | p |
| *Patients characteristics* |  |  |  |  |  |
| Age (years) | 80.86±5.71 | 80.72±5.59 | 80.61±6.13 | 81.17±5.48 | 0.403 |
| Male | 785/1797 (43.68%) | 206/491 (41.95%) | 210/438 (47.94%) | 220/463 (47.52%) | 0.118 |
| Body Mass Index (kg/m²) | 27.34±4.73 | 27.34±4.77 | 27.20±4.32 | 27.48±5.07 | 0.802 |
| Hypertension | 1690/1785 (94.68%) | 453/486 (93.21%) | 409/437 (93.59%) | 433/457 (94.75%) | 0.597 |
| Diabetes mellitus | 577/1788 (32.27%) | 160/488 (32.79%) | 149/437 (34.10%) | 154/458 (33.62%) | 0.912 |
| Insulin | 238/1759 (13.53%) | 65/483 (13.46%) | 71/424 (16.74%) | 72/447 (16.11%) | 0.339 |
| Dyslipidemia | 1170/1786 (65.51%) | 320/487 (65.71%) | 296/437 (67.73%) | 304/457 (66.52%) | 0.807 |
| Smoking | 122/1744 (6.99%) | 40/483 (8.28%) | 23/421 (5.46%) | 28/436 (6.42%) | 0.227 |
| Anemia | 977/1782 (54.83%) | 254/490 (51.84%) | 239/435 (54.94%) | 267/459 (58.17%) | 0.147 |
| COPD | 452/1786 (25.31%) | 122/487 (25.01%) | 111/437 (25.40%) | 138/458 (30.13%) | 0.151 |
| Neurological dysfunction | 146/1759 (8.30%) | 36/483 (7.45%) | 35/427 (8.20%) | 35/451 (7.76%) | 0.916 |
| Severe liver disease | 41/1783 (2.30%) | 13/488 (%) | 10/435 (%) | 10/457 (%) | 0.882 |
| PAD | 421/1758 (23.95%) | 128/483 (26.50%) | 90/428 (21.03%) | 109/451 (24.17%) | 0.155 |
| Carotid stenosis ≥50% | 45/1354 (3.23%) | 6/297 (%) | 10/338 (%) | 15/345 (%) | 0.235 |
| Critical preoperative state | 66/1779 (3.71%) | 18/485 (3.71%) | 23/435 (5.29%) | 18/456 (3.95%) | 0.454 |
| CAD history | 448/1784 (25.11%) | 125/488 (25.61%) | 109/436 (25.00%) | 130/456 (28.51%) | 0.441 |
| Prior myocardial infarction | 255/1786 (14.28%) | 76/488 (15.57%) | 64/436 (14.68%) | 86/457 (18.82%) | 0.208 |
| Prior cardiac surgery | 253/1787 (14.16%) | 68/489 (13.91%) | 68/437 (15.56%) | 72/458 (15.72%) | 0.687 |
| Prior myocardial revascularization | 406/1791 (22.68%) | 119/489 (24.33%) | 95/437 (21.74%) | 118/459 (25.71%) | 0.370 |
| PCI | 248/1791 (13.85%) | 76/489 (15.54%) | 48/437 (10.98%) | 74/459 (16.12%) | 0.056 |
| CABG | 92/1791 (5.14%) | 28/489 (5.73%) | 27/437 (6.18%) | 26/459 (5.66%) | 0.938 |
| PCI + CABG | 66/1791 (3.69%) | 15/489 (3.07%) | 20/437 (4.58%) | 18/459 (3.92%) | 0.486 |
| Myocardial revascularization for TAVR | 271/1791 (15.13%) | 67/486 (13.79%) | 54/437 (12.36%) | 65/463 (14.04%) | 0.728 |
| PCI | 266/1791 (14.85%) | 66/486 (13.58%) | 53/437 (12.13%) | 62/463 (13.39%) | 0.781 |
| CABG | 4/1791 (0.22%) | 1/486 (0.21%) | 1/437 (0.23%) | 2/463 (0.43%) | 0.778 |
| PCI + CABG | 1/1791 (0.05%) | 0/486 (0.00%) | 0/437 (0.00%) | 1/463 (0.22%) | 0.369 |
| Residual significant CAD | 214/1782 (12.01%) | 60/483 (12.42%) | 48/437 (12.34%) | 49/459 (10.67%) | 0.666 |
| during TAVR |  |  |  |  |  |
| Prior PM/ICD/CRT implantation | 222/1772 (12.53%) | 57/485 (11.75%) | 49/433 (11.32%) | 62/454 (13.66%) | 0.522 |
| NYHA functional class III-IV | 1475/1786 (82.59%) | 402/487 (82.55%) | 355/437 (81.24%) | 356/457 (77.90%) | 0.181 |
| CKD | 743/1797 (41.35%) | 212/491 (43.18%) | 178/438 (40.64%) | 200/463 (43.20%) | 0.671 |
| *Electrocardiography* |  |  |  |  |  |
| Sinus rhythm | 1462/1788 (81.77%) | 390/488 (79.92%) | 354/437 (81.01%) | 383/458 (83.62%) | 0.325 |
| Atrial fibrillation / flutter | 326/1788 (18.23%) | 98/488 (20.08%) | 83/437 (18.99%) | 75/458 (16.38%) | 0.325 |
| PM-induced rhythm | 94/1788 (5.26%) | 31/488 (6.35%) | 24/437 (5.49%) | 24/458 (5.24%) | 0.741 |
| *Echocardiography* |  |  |  |  |  |
| LVEF (%) | 53.34±10.21 | 54.22±10.81 | 52.88±10.47 | 52.33±9.77 | <0.001 |
| Maximum aortic gradient (mmHg) | 75.67±21.22 | 73.43±20.45 | 77.38±20.00 | 76.33±22.69 | 0.032 |
| Mean aortic gradient (mmHg) | 46.40±14.33 | 45.27±14.29 | 47.21±13.23 | 46.92±14.93 | 0.074 |
| Moderate-to-severe mitral regurgitation | 679/1678 (40.46%) | 175/455 (38.46%) | 179/407 (43.98%) | 194/429 (45.22%) | 0.095 |
| Pulmonary arterial systolic pressure (mmHg) | 40.22±13.37 | 40.23±12.47 | 39.72±12.84 | 40.07±12.83 | 0.834 |
| *Mortality risk scores* |  |  |  |  |  |
| Logistic EuroSCORE | 16.14±12.31 | 16.04±12.76 | 15.94±12.18 | 17.49±13.55 | 0.068 |
| EuroSCORE II | 5.79±12.75 | 5.61±5.84 | 5.26±5.35 | 6.04±6.79 | 0.283 |
| STS-PROM | 4.60±3.55 | 4.70±3.60 | 4.47±3.45 | 4.96±4.32 | 0.077 |
| STS-PROM≥8 | 176/1779 (9.90%) | 45/486 (9.26%) | 44/435 (10.11%) | 60/453 (13.24%) | 0.122 |

COPD = chronic obstructive pulmonary disease; PAD = peripheral artery disease; CAD = coronary artery disease; PCI = percutaneous coronary intervention; CABG = coronary artery by-pass grafting; TAVR = transcatheter aortic valve replacement; PM = pacemaker; ICD = implantable cardioverter-defibrillator; CRT = cardiac resynchronization therapy; NYHA = New York Heart Association; CKD = chronic kidney disease; LVEF = left ventricular ejection fraction; EuroSCORE = european system for cardiac operative risk evaluation; STS-PROM = Society of Thoracic Surgery predictive risk of mortality.

No statistically significant differences were found in terms of preprocedural characteristics like patients’ characteristics, previous cardiovascular history, comorbidities and mortality risk scores. Only echocardiographics parameters like left ventricular ejection fraction (p<0.001) and maximum aortic gradient (p = 0.032) were significantly different between the three groups.

## 3.2. Procedural characteristics

All procedural, post-procedural data and outcomes are shown in Table 2.

**Table 2.** Procedural features and outcomes according to FT tertiles.

| Variable | All | Fluoroscopy Time |  |  |  |
| --- | --- | --- | --- | --- | --- |
|  |  | 1<sup>st</sup> | 2<sup>nd</sup> | 3<sup>rd</sup> | p |
| *Procedural details* |  |  |  |  |  |
| Transfemoral access route | 1703/1797 (94.77%) | 444/491 (90.43%) | 416/438 (94.98%) | 440/463 (95.03%) | 0.005 |
| Other access routes | 94/1797 (5.23%) | 47/491 (9.57%) | 22/438 (5.02%) | 23/463 (4.97%) | 0.005 |
| Trans-subclavian | 27/1797 (1.57%) | 9/491 (1.83%) | 6/438 (1.37%) | 12/463 (2.59%) | 0.404 |
| transapical | 57/1797 (3.17%) | 36/491 (7.33%) | 10/438 (2.28%) | 9/463 (1.94%) | <0.001 |
| direct aortic | 8/1797 (0.44%) | 1/491 (0.20%) | 6/438 (1.37%) | 1/463 (0.22%) | 0.029 |
| Orotracheal intubation | 233/1796 (12.97%) | 73/491 (14.87%) | 66/437 (15.10%) | 90/463 (19.44%) | 0.107 |
| Valve-in-valve | 73/1794 (4.07%) | 16/491 (3.26%) | 20/437 (4.58%) | 27/461 (5.86%) | 0.157 |
| Pre-dilation | 827/1784 (46.36%) | 186/489 (38.04%) | 248/436 (56.88%) | 282/456 (61.84%) | <0.001 |
| Valve kind |  |  |  |  |  |
| balloon-expandable | 551/1797 (30.66%) | 190/491 (38.70%) | 153/438 (34.93%) | 130/463 (28.08%) | 0.002 |
| self-expandable | 1124/1797 (62.55%) | 266/491 (54.17%) | 248/438 (56.62%) | 294/463 (63.50%) | 0.011 |
| others | 122/1797 (6.79%) | 35/491 (7.13%) | 37/438 (8.45%) | 39/463 (8.42%) | 0.691 |
| Valve Size >26 mm | 722/1793 (40.27%) | 175/489 (35.79%) | 166/438 (37.90%) | 200/461 (43.38%) | 0.048 |
| Post-dilation | 479/1795 (26.68%) | 97/491 (19.76%) | 112/436 (25.69%) | 145/463 (31.32%) | <0.001 |
| CM volume (mL) | 149.97±76.36 | 130.431±54.03 | 161.14±67.22 | 197.61±96.94 | <0.001 |
| Radiation Dose (mGY) | 1366.18±1241.57 | 1070.68±1051.40 | 1381.24±1070.11 | 2112.15±1748.60 | <0.001 |
| **Complications and outcomes (VARC-3)** |  |  |  |  |  |
| AKI | 272/1714 (15.87%) | 70/472 (14.83%) | 50/420 (11.90%) | 72/433 (16.63%) | 0.142 |
| CVVH | 41/1730 (2.37%) | 10/476 (2.10%) | 8/420 (1.90%) | 15/438 (3.42%) | 0.288 |
| Chronic hemodialysis | 9/1667 (0.54%) | 3/460 (0.65%) | 2/409 (0.49%) | 4/423 (0.95%) | 0.724 |
| Bleeding (VARC-3) | 588/1399 (48.03%) | 124/343 (36.15%) | 139/313 (44.41%) | 214/362 (59.12%) | <0.001 |
| Type 1 | 192/ (13.72%) | 34/343 (9.91%) | 55/313 (17.57%) | 72/362 (19.89%) | <0.001 |
| Type 2 | 307/1399 (21.94%) | 67/343 (19.53%) | 67/313 (21.41%) | 102/362 (28.18%) | 0.017 |
| Type 3-5 | 89/1399 (6.36%) | 23/343 (6.71%) | 17/313 (5.43%) | 40/362 (11.05%) | 0.016 |
| BARC≥3 | 561/1773 (32.37%) | 124/477 (26.00%) | 117/424 (27.59%) | 208/449 (46.32%) | <0.001 |
| Need of transfusion | 298/1721 (17.31%) | 61/475 (12.84%) | 65/419 (15.51%) | 108/442 (24.43%) | <0.001 |
| 1 unit | 140/1721 (8.13%) | 30/475 (6.32%) | 31/419 (7.40%) | 52/442 (11.76%) | 0.008 |
| 2 units | 106/1721 (6.16%) | 23/475 (4.84%) | 23/419 (5.49%) | 38/442 (8.60%) | 0.046 |
| >2 units | 52/1721 (3.02%) | 8/475 (1.68%) | 11/419 (2.62%) | 18/442 (4.07%) | 0.086 |
| Vascular complications | 286/1765 (16.20%) | 59/484 (12.19%) | 57/436 (13.07%) | 118/455 (25.93%) | <0.001 |
| minor | 170/1765 (9.63%) | 41/484 (8.47%) | 34/436 (7.80%) | 65/455 (14.29%) | 0.002 |
| major | 116/1765 (6.57%) | 18/484 (3.72%) | 23/436 (5.27%) | 53/455 (11.65%) | <0.001 |
| Access-site related vascular complications | 224/339 (66.08%) | 22/69 (31.88%) | 43/86 (50.00%) | 47/137 (34.31%) | 0.029 |
| PCD failure | 101/1556 (6.49%) | 20/404 (4.72%) | 22/373 (5.90%) | 40/387 (10.34%) | 0.005 |
| At least moderate residual aortic regurgitation | 129/1562 (8.26%) | 26/412 (6.31%) | 30/377 (7.96%) | 38/398 (9.55%) | 0.223 |
| Permanent PM implantation | 226/1572 (14.38%) | 51/419 (12.17%) | 64/484 (16.67%) | 60/381 (15.75%) | 0.163 |
| ECM/cardiac arrest | 66/1713 (3.85%) | 15/473 (3.17%) | 7/425 (1.65%) | 34/440 (7.73%) | <0.001 |
| New-onset atrial fibrillation/flutter | 124/1412 (8.78%) | 33/375 (8.80%) | 25/343 (7.29%) | 33/376 (8.78%) | 0.707 |
| Acute myocardial infarction | 19/1774 (1.07%) | 2/486 (0.41%) | 3/437 (0.69%) | 11/454 (2.42%) | 0.009 |
| Stroke/TIA | 34/1773 (1.92%) | 6/487 (1.23%) | 8/437 (1.83%) | 7/453 (1.54%) | 0.759 |
| Hospital length of stay (days) | 5.73±9.63 | 4.99±3.52 | 5.53±3.93 | 6.34±4.46 | <0.001 |
| Hospital length of stay>5days | 627/1722 (36.41%) | 139/472 (29.45%) | 160/424 (37.74%) | 194/431 (45.01%) | <0.001 |
| Technical success | 1612/1752 (92.01%) | 456/480 (95.00%) | 406/431 (94.20%) | 382/453 (84.33%) | <0.001 |
| Device success | 1562/1764 (88.55%) | 442/485 (91.13%) | 383/431 (88.86%) | 385/451 (85.37%) | 0.021 |
| Periprocedural mortality | 43/1734 (2.48%) | 10/475 (2.10%) | 10/423 (2.36%) | 19/452 (4.21%) | 0.118 |
| Mortality at one year (F-U) | 59/1686 (3.50%) | 19/465 (4.09%) | 10/413 (2.42%) | 20/432 (4.63%) | 0.212 |
| Early safety absence (VARC-2) | 222/1732 (12.82%) | 42/477 (8.80%) | 43/426 (10.09%) | 87/449 (19.38%) | <0.001 |
| Early safety absence (VARC-3) | 559/1732 (32.27%) | 141/477 (29.56%) | 115/426 (26.99%) | 161/449 (35.86%) | 0.013 |
| Post-procedural complications (VARC-2 and VARC-3) | 427/839 (50.89%) | 91/203 (44.83%) | 100/188 (53.19%) | 146/250 (58.40%) | 0.016 |
| Complication time-delay (days) from TAVR | 3.44±39.39 | 3.13±18.55 | 6.99±79.80 | 2.55±11.97 | 0.049 |

AKI = acute kidney injury; CVVH = continuous venovenous haemofiltration; PCD = percutaneous closure device; ECM = external cardiac massage; TIA = transient ischemic attack; F-U = follow up; VARC = Valve Academic Research Consortium

Some procedural details, such as transfemoral access (p = 0.005), predilatation (p<0.001), self-expandable bioprosthesis (p = 0.011), valve size >26 mm (p = 0048), postdi-latation (p<0.001), contrast mean (CM) amount (p<0.001) and RD (p<0.001) were significantly associated with FT. With respect to complications according to VARC-3 criteria, FT was significantly associated with bleedings (p<0.001), transfusions (p<0.001), vascular complications (p<0.001), percutaneous closure device failure (p = 0.005), cardiac arrest during the procedure (p<0.001) and acute myocardial infarction (p = 0.009). FT resulted also significantly linked (p=0.016) with post-procedural complications and patients in the longest FT group experienced a complication earlier than those with shorter FT (p=0.049). Moreover, longer hospitalizations were significantly associated with higher FT during the TAVR procedure (p<0.001).

Furthermore, figure 1 shows the variation of FT and RD after splitting the population into tertiles according to the period of enrollment.

**Figure 1.**
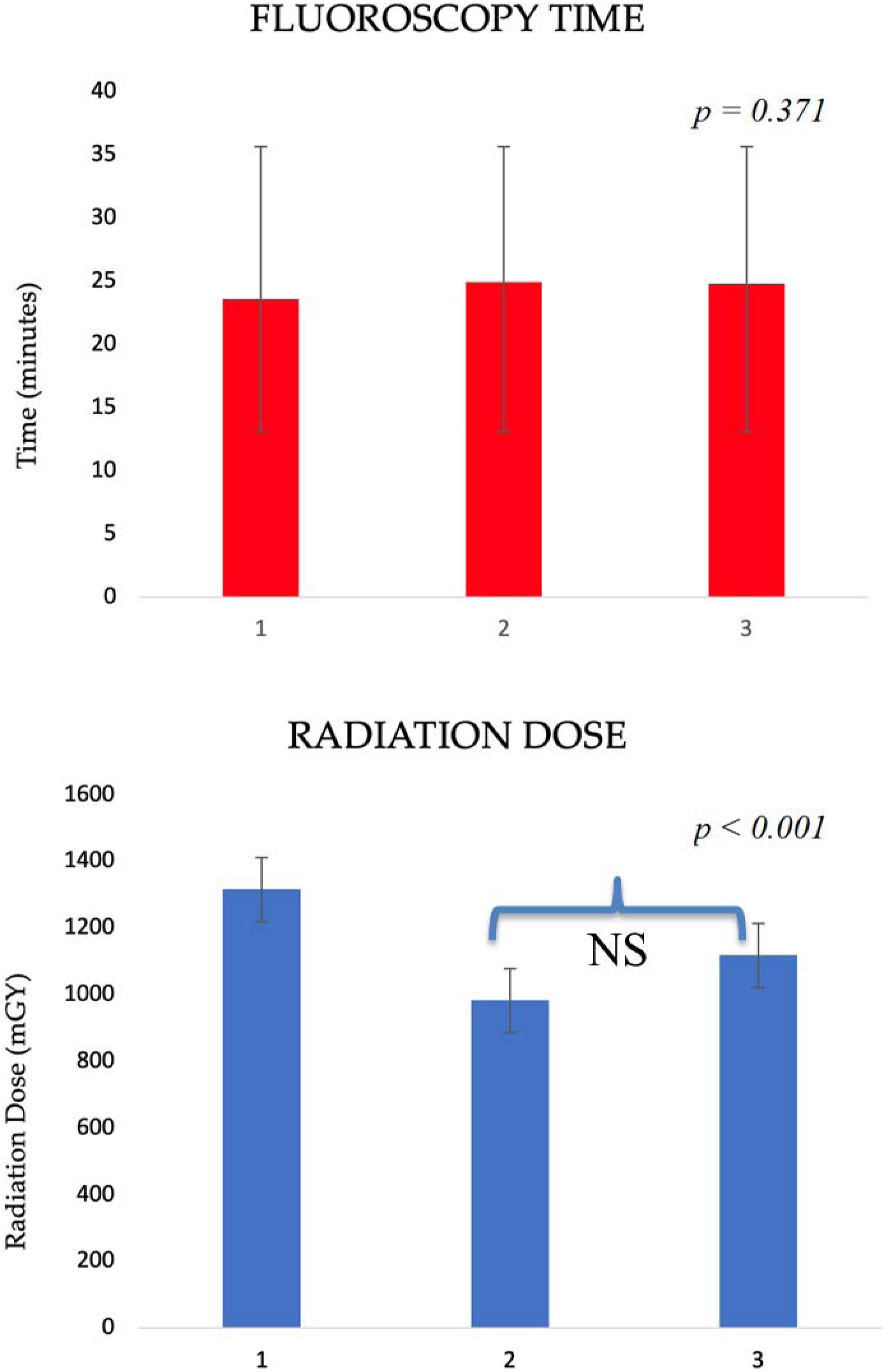
TAVR learning curve. Fluoroscopy time and Radiation dose (mean and standard deviation) according to TAVR enrollment time tertiles (1st tertile: 598 patients from April 2011 to September 2017; 2nd tertile: 600 patients from October 2017 to November 2020; 3rd tertile:

There was no significant difference of FT across the tertiles of enrollment-time (23.54±15.41; 24.92±18.13; 24.74±12.01 min; p = 0.371). On the other hand, there is a significant variation of RD that spanned along the study time (p<0.001). RD significantly decreased between first and second enrolling time tertile, while the slight RD increase between second and third tertiles resulted not significant after pairwise comparisons (1143.96±82.72 vs 1449.70±73.23 mGY; p=0.175).

## 3.3. Outcomes

Table 2 also shows outcomes incidence and its relationship with FT. Concerning outcomes according to VARC-3 criteria, higher FT was significantly associated with lower TS and DS (p<0.001 and p = 0.021 respectively) and higher absence of ES (p = 0.013). Also considering VARC-2 criteria, the absence of ES was significantly associated with the fluoroscopy time (p<0.001).

Furthermore, table 3 describes the relation between FT and outcomes, intended as TS, DS and ES according to VARC-3 criteria and ES according to VARC-2 criteria, after PSM. PSM was performed including, as covariates, all those variables that were considered to be likely to influence FT: pre-dilatation, post-dilatation, intraprocedural complications (according each VARC-2 or -3 outcome), self-expandable valve implantation, pre-TAVR ejection fraction, pre-TAVR maximum transaortic gradient, other vascular access than femoral one. After PSM, higher FT was still significantly related with the absence of TS (p = 0.001), DS (p < 0.001) and ES (p = 0.035), according to VARC-3 criteria, and the absence of ES (p = 0.046) according to VARC-2 criteria.

**Table 3.** Relationship between fluoroscopy time and outcomes before and after propensity score matching (PSM)

| Variable | Early Safety (VARC-3) |  | p | T-statistic<br>Yes |
| --- | --- | --- | --- | --- |
|  | Yes | No |  |  |
| Fluoroscopy Time min (unmatched) | 23.58±1.80 | 28.23±1.80 | <0.001 | 23.58±1.80 |
| Fluoroscopy Time min (matched after PSM*) | 24.09±3.17 | 28.23±3.17 | 0.035 | 24.09±3.17 |
|  | Device Success (VARC-3) |  | p |  |
|  | Yes | No |  |  |
| Fluoroscopy Time min (unmatched) | 23.70±1.93 | 31.95±1.93 | <0.001 | 4.27 |
| Fluoroscopy Time min (matched after PSM*) | 22.83±2.76 | 32.23±2.76 | 0.007 | 3.41 |
|  | Technical Success |  | p |  |
|  | Yes | No |  |  |
| Fluoroscopy Time min (unmatched) | 23.82±2.32 | 37.99±2.32 | <0.001 | 2.58 |
| Fluoroscopy Time min (matched after PSM*) | 22.01±4.23 | 37.99±4.23 | 0.001 | 1.90 |
|  | Early Safety (VARC-2) |  | p |  |
|  | Yes | No |  |  |
| Fluoroscopy Time min (unmatched) | 33.72±2.05 | 23.72±3.05 | <0.001 | 4.87 |
| Fluoroscopy Time min (matched after PSM*) | 33.72±3.10 | 22.11±3.10 | 0.046 | 3.74 |

Moreover, the ROC analysis showed a significant correlation between FT and these outcomes (Table 4, Figure 2). Nevertheless, based on the AUC of the cut-off values established with the highest Youden’s indexes, a good performance was demonstrated only in detecting TS (Cut-off 27.8±0.04 min) according to VARC-3 criteria and ES (Cut-off 30.1±0.03 min) according to VARC-2 criteria (ES-VARC-3: AUC 0.545, 95% CI 0.518–0.571, sensitivity 29.5%, specificity 80%, p = 0.008; DS-VARC-3: AUC 0.608, 95% CI 0.581–0.633, sensitivity 60.56%, specificity 57.89%, p < 0.001, TS-VARC-3: AUC 0.680, 95% CI 0.654–0.704, sensitivity 54.17%, specificity 76.21%, p <0.001; ES-VARC-2: AUC 0.628, 95% CI 0.601–0.654, sensitivity 41.28%, specificity 81.53%, p <0.001).

**Figure 2.**
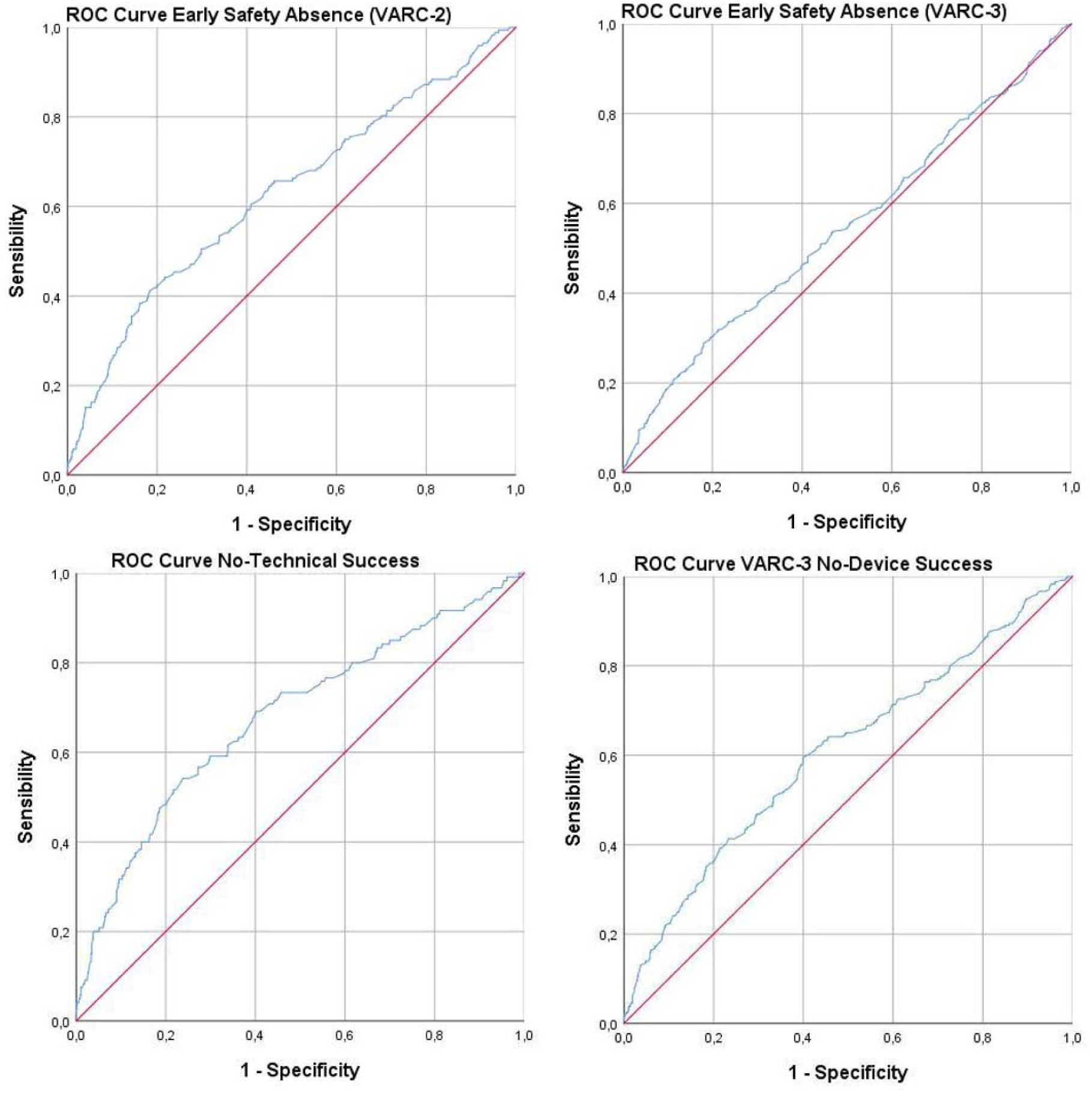
Absence of ES, DS and TS according to FT: ROC curve analysis.9 patients from December 2020 to april 2023). NS: not significant.

**Table 4.**
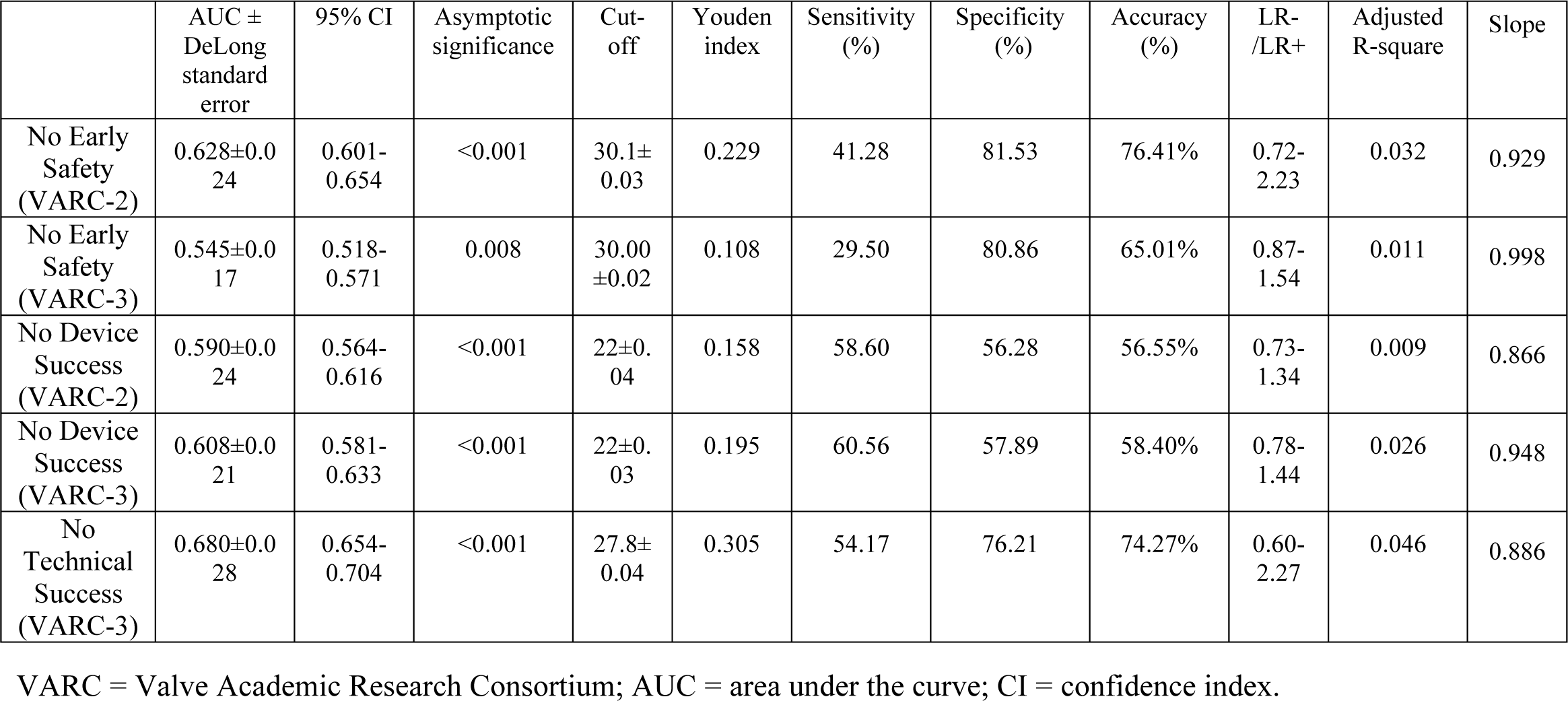
ROC analysis of VARC-2 and VARC-3 outcomes according to FT.

### 4. Discussion

The main findings of our study can be summarized as follows: 1) longer FT during TAVR is related to more complex procedure; 2) short-term outcomes after TAVR are related with FT and this association persists after PSM; 3) the cut-offs identified after ROC analysis have sufficient accuracy to detect VARC-3 TS and VARC-2 ES; 4) the variation over time of FT is non significantly related with TAVR learning curve.

Our study is the first large TAVR cohort in which has been investigated the relationship between FT and short-term outcomes after the last updated consensus document VARC-3 [22]. Previous studies have demonstrated that prolonged FT is associated with more complex lesions treated in percutaneous coronary intervention [23-25]. In our analysis a longer FT is associated with transfemoral access approach and a more challenging procedure: need of pre-dilatation and post-dilatation, self-expandable valve implantation, higher CM amount used and higher radation dose produced during the procedure. However, the higher length of FT when transfemoral access is used instead of a surgical approach (like trans-apical and trans-sublacvian) is easily explained by the fact that in the first case the management of the access is completely cine-fluoroscopic guided in the majority of procedures.

To date, no study analyzed the relationship between FT and short-term outcomes (according VARC-2 and -3 definitions) in TAVR. Only radiation exposure during the procedure has been investigated and has been shown to be comparable to percutaneous coronary intervention of moderate complexity [18-20]. Unlike radiation exposure, FT is independent by biometric parameters such as BMI. Interestingly, in our study the absence of all short-term outcomes (VARC-3 TS, DS and ES and VARC-2 ES) is significantly associated with longer FT and this association persists after PSM including as covariates all variables that could influence the length of the procedure, including intra-procedural complications. FT appears to be an independent predictor of short-term TAVR-related post-procedural complications. This could be explained by the fact that in interventional cardiology the less procedures last, the fewer are the complications and consequently the better are the outcomes.

However, although the ROC analysis showed a significant correlation between the FT and these outcomes, the identified cut-offs do not have adequate diagnostic accuracy in adjudicating DS and ES according to VARC-3 criteria, due to the fact that their AUC was never above 0.6. Conversely, the cut-offs of 27.8±0.04 min for TS according to VARC-3 and of 30.1±0.03 for ES according to VARC-2 have adequate diagnostic accuracy. These values show that when the FT lasts more than 30 minutes, it is more likely that the patient might have to experience technical failure at the exit of the catheterization laboratory or might have short-term complications according VARC-2 and VARC-3 definitions.

The reason why this cut-off is able to predict the absence of ES-VARC-2 instead of ES VARC-3 can properly be explained by the possible limits of new VARC-3 criteria: the more complications are included in this composite endpoint, the more its diagnostic performance will decrease. Indeed, as previously reported, even the accuracy of the mortality risk score is lower with VARC-3 than with VARC-2 criteria [26].

Finally, looking at TAVR learning curve (Figure 1), there was no significant variation in FT, while a significant reduction of RD administered during the period of enrollment resulted significant only between the first and the second tertile of en-rollment period. This finding could be explained by the fact that the first tertile of enrollment, covering about 7 years, spanned for a longer time than the second and the third ones. So the first tertile of enrollment comprehends the most of the learning curve. The RD reduction is maybe related to technological advancements of the newer radiological angiographers. Another explanation is that the majority of invasive coronary angiography were performed simultaneously with TAVR in the third tertile of enrollment-time. Thus, even though the operators improved their skills there wasn’t a decrease of both FT and RD between the second and third period of TAVR enrollment due to the fact that more procedures were performed at the same time. However our data showed no significative increase in the revascularization rate during TAVR according to FT tertiles, so an indirect linkage between FT and myocardial injury related to myocardial revascularization during TAVR could be excluded.

### 5. Limitations

Although it was obtained from a prospectively collected database, this is an unspecified post-hoc analysis. Therefore, we cannot exclude that potential confounding factors not considered in the model may have influenced the results. The effect of a learning curve and changes in treatment strategy is also heterogeneous, as the study spanned more than a decade. Furthermore, we believe that aspects of management not controlled or specified may have been a source of bias. Finally, an independent committee did not adjudicate all clinical events that were site-referred.

### 6. Conclusions

FT is an easily available parameter during TAVR. This is the first study that evaluated the relationship between FT and short-term outcomes after TAVR. Longer FT is related to peri-procedural and short-term outcomes after TAVR, expecially in more complex procedures. This relation is independent from intra-procedural complications after PSM analysis. A FT duration of more than 30 minutes has an adequate accuracy in identifying VARC-3 technical failure and absence of VARC-2 ES. Defining ES according to the most recent VARC-3 consensus document invalidates the accuracy of this relation, probably because of the inclusion of additional frequent adverse events such as permanent PM implantation.

## Acknowledgments

Francesco Spione has been supported by a research grant provided by the Cardiopath PhD program.

## Acknowledgment of grant support

none.

## Data Availability Statement

Data are accessible with the article (raw data are available upon individual request).

## Notes

**Funding:** This research received no external funding.

### Competing Interest Statement

Gaetano Contegiacomo serves as transcatheter heart valve proctor for Abbott; Fortunato Iacovelli and Francesco Spione directly received speaker fees from General Electric Healthcare; the remaining authors have no conflicts of interest to declare.

### Clinical Trial

The study was conducted in accordance with the Declara-tion of Helsinki, and approved by the Independent Ethics Committee of the Policlinico University Hospital of Bari, Italy (study number 6244, protocol code 0030669 and date of approval 04th of March 2020).

### Funding Statement

This research received no external funding.

## REFERENCES

1. Smith, C.R.; Leon, M.B.; Mack, M.J.; Miller, D.C.; Moses, J.W.; Svensson, L.G.; Tuzcu, E.M.; Webb, J.G.; Fontana, G.P.; Makkar, R.R.;, et al. Transcatheter versus surgical aortic-valve replacement in high-risk patients. N Engl J Med 2011, 364, 2187–2198, doi:10.1056/NEJMoa1103510.

2. Lefèvre, T.; Kappetein, A.P.; Wolner, E.; Nataf, P.; Thomas, M.; Schächinger, V.; De Bruyne, B.; Eltchaninoff, H.; Thielmann, M.; Himbert, D.;, et al. One year follow-up of the multi-centre European PARTNER transcatheter heart valve study. Eur Heart J 2011, 32, 148–157, doi:10.1093/eurheartj/ehq427.

3. Eltchaninoff, H.; Prat, A.; Gilard, M.; Leguerrier, A.; Blanchard, D.; Fournial, G.; Iung, B.; Donzeau-Gouge, P.; Tribouilloy, C.; Debrux, J.L.;, et al. Transcatheter aortic valve implantation: early results of the FRANCE (FRench Aortic National CoreValve and Edwards) registry. Eur Heart J 2011, 32, 191–197, doi:10.1093/eurheartj/ehq261.

4. Thomas, M.; Schymik, G.; Walther, T.; Himbert, D.; Lefèvre, T.; Treede, H.; Eggebrecht, H.; Rubino, P.; Michev, I.; Lange, R.;, et al. Thirty-day results of the SAPIEN aortic Bioprosthesis European Outcome (SOURCE) Registry: A European registry of transcatheter aortic valve implantation using the Edwards SAPIEN valve. Circulation 2010, 122, 62–69, doi:10.1161/circulationaha.109.907402.

5. Rodés-Cabau, J.; Webb, J.G.; Cheung, A.; Ye, J.; Dumont, E.; Feindel, C.M.; Osten, M.; Natarajan, M.K.; Velianou, J.L.; Martucci, G.;, et al. Transcatheter aortic valve implantation for the treatment of severe symptomatic aortic stenosis in patients at very high or prohibitive surgical risk: acute and late outcomes of the multicenter Canadian experience. J Am Coll Cardiol 2010, 55, 1080–1090, doi:10.1016/j.jacc.2009.12.014.

6. Petronio, A.S.; De Carlo, M.; Bedogni, F.; Marzocchi, A.; Klugmann, S.; Maisano, F.; Ramondo, A.; Ussia, G.P.; Ettori, F.; Poli, A.;, et al. Safety and efficacy of the subclavian approach for transcatheter aortic valve implantation with the CoreValve revalving system. Circ Cardiovasc Interv 2010, 3, 359–366, doi:10.1161/circinterventions.109.930453.

7. Leon, M.B.; Smith, C.R.; Mack, M.; Miller, D.C.; Moses, J.W.; Svensson, L.G.; Tuzcu, E.M.; Webb, J.G.; Fontana, G.P.; Makkar, R.R.;, et al. Transcatheter aortic-valve implantation for aortic stenosis in patients who cannot undergo surgery. N Engl J Med 2010, 363, 1597–1607, doi:10.1056/NEJMoa1008232.

8. Himbert, D.; Descoutures, F.; Al-Attar, N.; Iung, B.; Ducrocq, G.; Détaint, D.; Brochet, E.; Messika-Zeitoun, D.; Francis, F.; Ibrahim, H.;, et al. Results of transfemoral or transapical aortic valve implantation following a uniform assessment in high-risk patients with aortic stenosis. J Am Coll Cardiol 2009, 54, 303–311, doi:10.1016/j.jacc.2009.04.032.

9. Piazza, N.; Grube, E.; Gerckens, U.; den Heijer, P.; Linke, A.; Luha, O.; Ramondo, A.; Ussia, G.; Wenaweser, P.; Windecker, S.;, et al. Procedural and 30-day outcomes following transcatheter aortic valve implantation using the third generation (18 Fr) corevalve revalving system: results from the multicentre, expanded evaluation registry 1-year following CE mark approval. EuroIntervention 2008, 4, 242–249, doi:10.4244/eijv4i2a43.

10. Grube, E.; Buellesfeld, L.; Mueller, R.; Sauren, B.; Zickmann, B.; Nair, D.; Beucher, H.; Felderhoff, T.; Iversen, S.; Gerckens, U. Progress and current status of percutaneous aortic valve replacement: results of three device generations of the CoreValve Revalving system. Circ Cardiovasc Interv 2008, 1, 167–175, doi:10.1161/circinterventions.108.819839.

11. Leon, M.B.; Smith, C.R.; Mack, M.J.; Makkar, R.R.; Svensson, L.G.; Kodali, S.K.; Thourani, V.H.; Tuzcu, E.M.; Miller, D.C.; Herrmann, H.C.;, et al. Transcatheter or Surgical Aortic-Valve Replacement in Intermediate-Risk Patients. N Engl J Med 2016, 374, 1609–1620, doi:10.1056/NEJMoa1514616.

12. Mack, M.J.; Leon, M.B.; Thourani, V.H.; Makkar, R.; Kodali, S.K.; Russo, M.; Kapadia, S.R.; Malaisrie, S.C.; Cohen, D.J.; Pibarot, P.;, et al. Transcatheter Aortic-Valve Replacement with a Balloon-Expandable Valve in Low-Risk Patients. N Engl J Med 2019, 380, 1695–1705, doi:10.1056/NEJMoa1814052.

13. D’Errigo, P.; Barbanti, M.; Ranucci, M.; Onorati, F.; Covello, R.D.; Rosato, S.; Tamburino, C.; Santini, F.; Santoro, G.; Seccareccia, F. Transcatheter aortic valve implantation versus surgical aortic valve replacement for severe aortic stenosis: results from an intermediate risk propensity-matched population of the Italian OBSERVANT study. Int J Cardiol 2013, 167, 1945–1952, doi:10.1016/j.ijcard.2012.05.028.

14. Thyregod, H.G.; Steinbrüchel, D.A.; Ihlemann, N.; Nissen, H.; Kjeldsen, B.J.; Petursson, P.; Chang, Y.; Franzen, O.W.; Engstrøm, T.; Clemmensen, P.;, et al. Transcatheter Versus Surgical Aortic Valve Replacement in Patients With Severe Aortic Valve Stenosis: 1-Year Results From the All-Comers NOTION Randomized Clinical Trial. J Am Coll Cardiol 2015, 65, 2184–2194, doi:10.1016/j.jacc.2015.03.014.

15. Popma, J.J.; Deeb, G.M.; Yakubov, S.J.; Mumtaz, M.; Gada, H.; O’Hair, D.; Bajwa, T.; Heiser, J.C.; Merhi, W.; Kleiman, N.S.;, et al. Transcatheter Aortic-Valve Replacement with a Self-Expanding Valve in Low-Risk Patients. N Engl J Med 2019, 380, 1706–1715, doi:10.1056/NEJMoa1816885.

16. Chambers, C.E.; Fetterly, K.A.; Holzer, R.; Lin, P.J.; Blankenship, J.C.; Balter, S.; Laskey, W.K. Radiation safety program for the cardiac catheterization laboratory. Catheter Cardiovasc Interv 2011, 77, 546–556, doi:10.1002/ccd.22867.

17. Little, M.P. Risks associated with ionizing radiation. Br Med Bull 2003, 68, 259–275, doi:10.1093/bmb/ldg031.

18. Sharma, D.; Ramsewak, A.; O’Conaire, S.; Manoharan, G.; Spence, M.S. Reducing radiation exposure during transcatheter aortic valve implantation (TAVI). Catheterization and Cardiovascular Interventions 2015, 85, 1256–1261, doi:https://doi.org/10.1002/ccd.25363.

19. Daneault, B.; Balter, S.; Kodali, S.K.; Williams, M.R.; Généreux, P.; Reiss, G.R.; Paradis, J.M.; Green, P.; Kirtane, A.J.; Smith, C.;, et al. Patient radiation exposure during transcatheter aortic valve replacement procedures. EuroIntervention 2012, 8, 679–684, doi:10.4244/eijv8i6a106.

20. Michel, J.M.; Hashorva, D.; Kretschmer, A.; Alvarez-Covarrubias, H.A.; Mayr, N.P.; Pellegrini, C.; Rheude, T.; Frangieh, A.H.; Giacoppo, D.; Kastrati, A.;, et al. Evaluation of a Low-Dose Radiation Protocol During Transcatheter Aortic Valve Implantation. Am J Cardiol 2021, 139, 71–78, doi:10.1016/j.amjcard.2020.10.035.

21. Kappetein, A.P.; Head, S.J.; Généreux, P.; Piazza, N.; van Mieghem, N.M.; Blackstone, E.H.; Brott, T.G.; Cohen, D.J.; Cutlip, D.E.; van Es, G.-A.;, et al. Updated standardized endpoint definitions for transcatheter aortic valve implantation: the Valve Academic Research Consortium-2 consensus document†. European Heart Journal 2012, 33, 2403–2418, doi:10.1093/eurheartj/ehs255.

22. Genereux, P.; Piazza, N.; Alu, M.C.; Nazif, T.; Hahn, R.T.; Pibarot, P.; Bax, J.J.; Leipsic, J.A.; Blanke, P.; Blackstone, E.H.;, et al. Valve Academic Research Consortium 3: updated endpoint definitions for aortic valve clinical research. Eur Heart J 2021, 42, 1825–1857, doi:10.1093/eurheartj/ehaa799.

23. Nikolsky, E.; Pucelikova, T.; Mehran, R.; Balter, S.; Kaufman, L.; Fahy, M.; Lansky, A.J.; Leon, M.B.; Moses, J.W.; Stone, G.W.;, et al. An evaluation of fluoroscopy time and correlation with outcomes after percutaneous coronary intervention. J Invasive Cardiol 2007, 19, 208–213.

24. Tajti, P.; Ayoub, M.; Nuehrenberg, T.; Ferenc, M.; Behnes, M.; Buettner, H.J.; Neumann, F.J.; Mashayekhi, K. Association of Prolonged Fluoroscopy Time with Procedural Success of Percutaneous Coronary Intervention for Stable Coronary Artery Disease with and without Chronic Total Occlusion. J Clin Med 2021, 10, doi:10.3390/jcm10071486.

25. Ishibashi, S.; Sakakura, K.; Asada, S.; Taniguchi, Y.; Yamamoto, K.; Tsukui, T.; Seguchi, M.; Jinnouchi, H.; Wada, H.; Fujita, H. Clinical Factors Associated with Long Fluoroscopy Time in Percutaneous Coronary Interventions to the Culprit Lesion of Non-ST-Segment Elevation Myocardial Infarction. Int Heart J 2021, 62, 282–289, doi:10.1536/ihj.20-634.

26. Iacovelli, F.; Loizzi, F.; Cafaro, A.; Burattini, O.; Salemme, L.; Cioppa, A.; Rizzo, F.; Palmitessa, C.; D’Alessandro, M.; De Feo, D.;, et al. Surgical Mortality Risk Scores in Transcatheter Aortic Valve Implantation: Is Their Early Predictive Value Still Strong? J. Cardiovasc. Dev. Dis. 2023, 10, doi:10.3390/jcdd10060244.

